# Current tobacco smoking and risk from COVID-19: results from a population symptom app in over 2.4 million people

**DOI:** 10.1101/2020.05.18.20105288

**Authors:** Nicholas S Hopkinson, Niccolò Rossi, Julia S El-Sayed Moustafa, Anthony A Laverty, Jennifer K Quint, Maxim B Freidin, Alessia Visconti, Benjamin Murray, Marc Modat, Sebastien Ourselin, Kerrin Small, Richard Davies, Jonathan Wolf, Tim D Spector, Claire J Steves, Mario Falchi

## Abstract

**Background:** The association between current tobacco smoking, the risk of developing COVID-19 and the severity of illness is an important information gap.

**Methods:** UK users of the COVID Symptom Study app provided baseline data including demographics, anthropometrics, smoking status and medical conditions, were asked to log symptoms daily from 24^th^ March 2020 to 23^rd^ April 2020. Participants reporting that they did not feel physically normal were taken through a series of questions, including 14 potential COVID-19 symptoms and any hospital attendance. The main study outcome was the association between current smoking and the development of “classic” symptoms of COVID-19 during the pandemic defined as fever, new persistent cough and breathlessness. The number of concurrent COVID-19 symptoms was used as a proxy for severity. In addition, association of subcutaneous adipose tissue expression of *ACE2*, both the receptor for SARS-CoV-2 and a potential mediator of disease severity, with smoking status was assessed in a subset of 541 twins from the TwinsUK cohort.

**Results:** Data were available on 2,401,982 participants, mean(SD) age 43.6(15.1) years, 63.3% female, overall smoking prevalence 11.0%. 834,437 (35%) participants reported being unwell and entered one or more symptoms. Current smokers were more likely to develop symptoms suggesting a diagnosis of COVID-19; classic symptoms adjusted OR[95%CI] 1.14[1.10 to 1.18]; >5 symptoms 1.29[1.26 to 1.31]; >10 symptoms 1.50[1.42 to 1.58]. Smoking was associated with reduced *ACE2* expression in adipose tissue (Beta(SE)=-0.395(0.149); p=7.01×10^-3^).

**Interpretation:** These data are consistent with smokers having an increased risk from COVID-19.

**Funding:** Zoe provided in kind support for all aspects of building, running and supporting the app and service to all users worldwide. The study was also supported by grants from the Wellcome Trust, UK Research and Innovation and British Heart Foundation.

**RESEARCH IN CONTEXT:** *Evidence before this study:* The interaction between current smoking and COVID-19 is unclear. Smoking is known to increase susceptibility to viral infections and appears to be associated with worse outcomes in people admitted to hospital with COVID-19. However, case series have reported relatively low levels of current smoking among individuals admitted to hospital with the condition, raising the possibility that smoking has a protective effect against the disease.

*Added value of this study:* Data from a large UK population who are users of a symptom reporting app during the pandemic supports the hypothesis that smokers are more likely to develop symptoms consistent with COVID-19 and that they have an increased symptom burden.

*Implications of all the available evidence:* These population data, combined with evidence of a worse outcome in smokers hospitalised with the condition, support the contention that smoking increases individual risk from COVID-19. Support to help people to quit smoking should therefore form part of efforts to deal with the pandemic.

## INTRODUCTION

Although in many people infection with SARS-CoV-2 causes no or only minor symptoms, in a proportion who develop COVID-19 there is progressive lung involvement with respiratory failure and widespread systemic consequences^1,2^.The risk of severe complications is higher in older people and those with long term medical conditions including cardiovascular disease, diabetes and COPD.^3-5^

Tobacco smoking is a significant risk factor for both viral and bacterial infections of the respiratory system^6,7^, with smokers five times as likely to develop influenza and twice as likely to have pneumonia^8^. It may therefore be an important factor worsening the impact of COVID-19. There is evidence from case series that smoking is associated with more severe disease, a greater risk of ITU admission and excess mortality in people with COVID-19 admitted to hospital^3,9-12^. However some reports have suggested that although smoking related disease is common in patients with COVID-19, current smoking rates in hospitalised patients are lower than would be expected from population smoking prevalence. While some studies have looked specifically at current smoking^10-12^ others combined current and ex-smokers^3,9^, and smoking prevalence in these samples suggest data may have been incomplete. A potential protective effect of nicotine has been suggested^13^.

It has been reported that smoking is associated with upregulation of ACE2^14-16^, the receptor for the SARS-CoV-2 virus^17^ in the lung, though a recent meta-analysis suggests divergent effects with upregulation in epithelial cells and downregulation in alveolar type 2 cells^18^. The situation is further complicated by the possibility that internalisation of ACE2 due to viral infection leads to unopposed ACEI activity and high angiotensin II levels, contributing to endothelial damage and the coagulopathy and microthrombosis seen in severe COVID-19^1,2^. Smoking itself causes vascular endothelial damage^19^, a prominent feature in the pathophysiology of severe COVID-19.

In order to establish the impact of current smoking more accurately, we analysed data from a population COVID-19 symptom reporting app, the COVID Symptom Study, developed by Zoe in collaboration with KCL and MGH. We hypothesised that current smokers would be at increased risk of developing COVID-19 symptoms and would experience a higher symptom burden. To provide additional mechanistic data we analysed the relationship between smoking and *ACE2* gene expression in adipose tissue samples from mono- and dizygotic twins from the TwinsUK cohort^20^ as well as investigating the relationship between *ACE2* expression in lung and adipose tissue in samples from the Genotype-Tissue Expression (GTEx) resource^21^.

## METHODS

### Data and analysis

Adult members of the public were invited to download the COVID Symptom Study App after launch on radio, TV and via social media. On first use, the app records self-reported location, age, and core health risk factors, including height, weight, smoking and common disease (*e.g*, diabetes, heart disease, lung disease) status, some medication use, as well as whether they thought that they had already had COVID-19^22^. With continued use, participants can provide daily updates on whether they have been tested for SARS-CoV-2 and if they “feel physically normal”. If not, they can record the presence of 14 COVID-19 related symptoms (*i.e*., abdominal pain, chest pain, delirium, diarrhoea, fatigue, fever, headache, hoarse voice, loss of smell, persistent cough, shortness of breath, skipped meals, sore throat, unusual muscle pains) and whether they had attended hospital. Fatigue and shortness of breath were graded by symptom severity, ranging from mild to severe.

The study has been approved by the King’s College London Research Ethics Committee REMAS ID 18210, review reference LRS-19/20-18210 and all subscribers provided informed consent.

### Study Population

The study population comprised individuals resident in the UK who had entered data via the app between 24th March and 23rd April 2020, meeting the following criteria: age 16 to 90 years; BMI (kg/m^2^) between 15 and 55. For this analysis we removed individuals with inconsistent SARS-CoV-2 test results (*i.e*., people reporting testing both positive and negative for SARS-CoV-2), and inconsistent assessments (*e.g*. where the body temperature, when logged, was outside the range of 35 to 43° C or where they reported feeling unwell but had no symptoms). We also excluded women who declared themselves to be pregnant (n=8,680).

We collected all symptoms declared by each participant during the period which they engaged with the app (median [IQR] number of days of 14[1-26]). While the response option for the majority of symptoms was yes/no, fatigue and shortness of breath were considered present if any level of severity was reported.

### Statistical analysis

Statistical analyses were carried out using R (v. 3.6.0). Association between smoking status and binary variables, (*i.e*., presence/absence of one or more symptoms, self-assessed COVID-19 status or results from testing for SARS-CoV-2) was carried out through multivariate logistic regression using the R *stats* package, and including age, sex and BMI as covariates.

The Bonferroni-adjusted threshold for statistical significance was calculated by dividing a conventional alpha value of 0.05 by the number of tested groups and the effective number of independent symptoms, which were estimated based on their pairwise polychoric correlation coefficients (*polycor* R package, v. 0.7-10), using the Li’s method^23^. The resulting p-value threshold was 0.05/4/11 = 1.14×10^-3^. Association p-values lower than this threshold were considered to be statistically significant.

In order to assess if smokers were characterised by specific patterns of co-occurring symptoms, we built distinct symptom correlation matrices in smokers and non-smokers based on the pairwise polychoric correlations between symptoms. Subsequently, we tested the correlation between the two matrices using the Mantel test, as implemented in the *vegan* R package (v. 2.5-6). The empirical p-value was given as the proportion of 1000 random permutations of the correlation matrices that had test statistic greater than or equal to the observed one.

We further explored the effect of smoking on COVID-19 severity among Standard Users (*i.e*., subjects neither tested for SARS-CoV-2 nor self-reporting prior suspected COVID-19 infection at the time of registration) by seeking association between smoking status and the development of a greater symptom burden. Tested logistic regression models included: i) presence of all classic COVID-19 related symptoms (*i.e*., fever, persistent cough and shortness of breath), ii) presence of more than 5 symptoms and iii) presence of more than 10 symptoms. Age, sex and BMI were included as covariates.

With continued use of the app, visits to hospitals were recorded through user updates on their location following suspected COVID-19 (*i.e*., “home”, “hospital”, or “back from hospital”). We treated hospital attendance as an additional proxy for COVID-19 symptom severity, and therefore tested by logistic regression its association with smoking in individuals testing positive for SARS-CoV-2. Those reporting being at the hospital or being back from the hospital at any login were considered as cases, whereas subjects who did not seek hospital care were treated as controls. Age, sex, BMI and presence of comorbidities (*i.e*., diabetes, lung disease, kidney disease, cancer and heart disease) or comorbidity-related treatments (*i.e*., blood pressure medications and immunosuppressants) were included as covariates in the model. As part of a sensitivity analysis, we also assessed the association between smoking status and hospital attendance (including age, sex and BMI as covariates) only in those subjects not reporting any comorbidities or treatments, in order to reduce the possible confounding effect of pre-existing health conditions.

### *ACE2* expression analyses

Subcutaneous adipose tissue gene expression, measured by RNASeq, was available in 541 female subjects who were participants in the TwinsUK cohort^20^. This included 54 smokers, 196 past smokers and 291 non-smokers. Association of *ACE2* expression in adipose tissue with smoking status was assessed using multivariate linear mixed effects models adjusting for RNASeq technical covariates, i.e. age, BMI, type 2 diabetes status, and family structure. Further details of these analyses are available in the *Online Supplement*.

### Assessment of *ACE2* expression in adipose tissue and lung samples from the GTEx resource

The data used for the analyses described in this manuscript were obtained from the GTEx Portal^24^ on 24/04/2020. Correlation of *ACE2* gene expression residuals in subcutaneous adipose tissue and lung, adjusted for age, sex and technical covariates, was assessed by Spearman’s correlation in R v 3.5.1 (See *Online Supplement* for further details).

## RESULTS

Overall, 2,401,982 UK residents, a majority female (63.3%), entered data via the app between 24th March 2020 and 22nd April 2020. Median (IQR) per-user number of login days was 14 (1 to 26). During this period, 834,437 individuals (35%) reported that they “did not feel physically normal” and entered at least one COVID-19 related symptom. The 2,401,982 participants were classified into the following categories: tested SARS-CoV-2 positive (SC2P; n=7,123); tested SARS-CoV-2 negative (SC2N; n=16,759); subjects self-reporting COVID-19 (SC2S; n=157,406) who believed, based on symptoms, that they had already had COVID-19 at the time of registration; the remaining 2,221,088 were defined as “Standard Users” (**Table 1**)

**Table 1.**
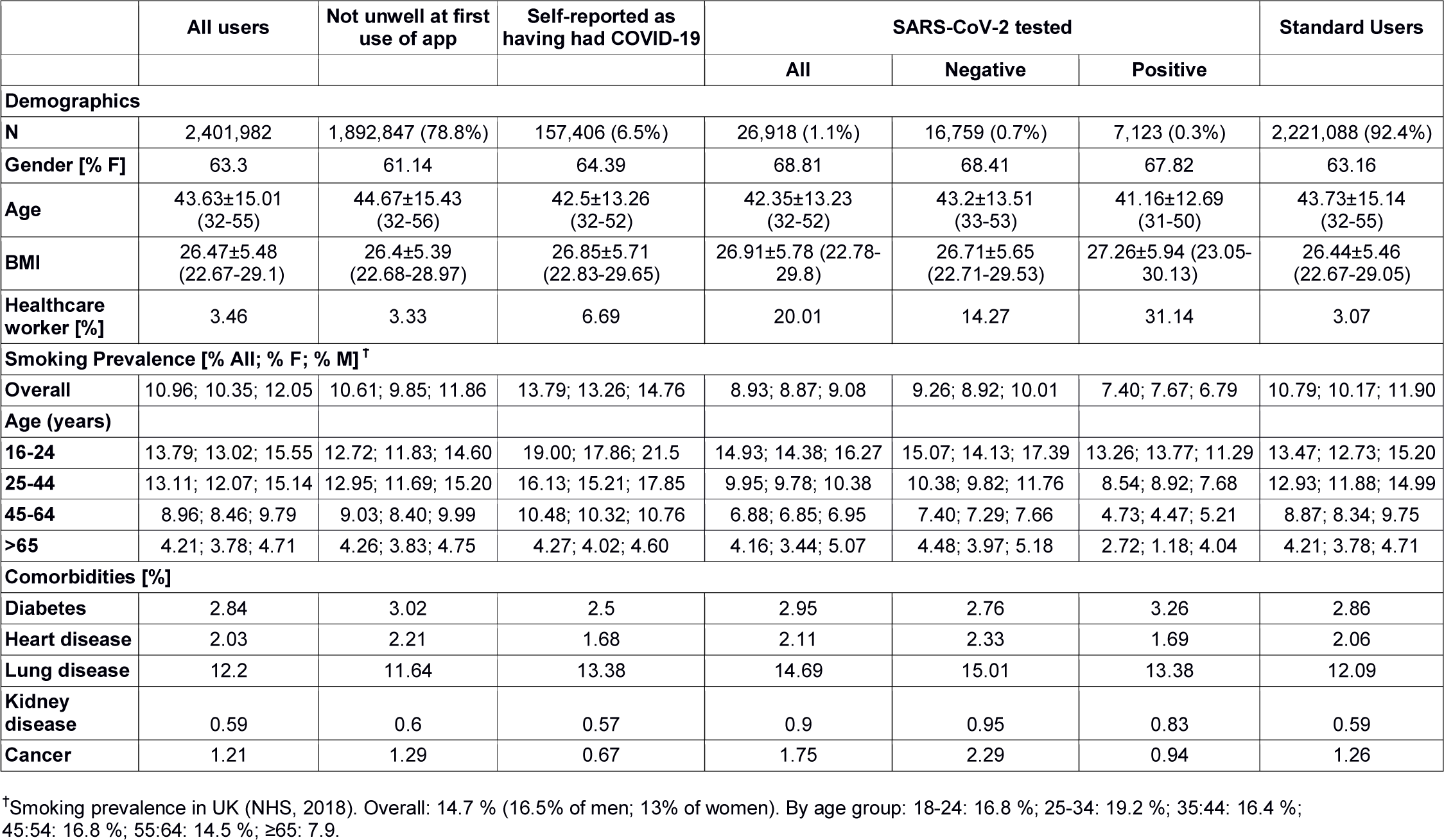
Characteristics of study subjects. Age and BMI are given as mean ± standard deviations and 1st, 3rd interquartile range (in brackets). Standard users refers to individuals who did not believe that they had already had COVID-19 when registering with the app and had not been tested for SARS-CoV-2.

The overall smoking rate in this sample was 11.0%. Smoking prevalence in the overall population and in any age and sex strata was lower compared to the general UK population (NHS report, 2018; **Table 1**). This difference may be due to sampling bias, reflecting the demographics of this sample, as smoking rates are lower in wealthier groups^25^ who may also have been more likely to be able to make use of a smartphone app. We noted a higher longitudinal drop-out rate for smokers, resulting in shorter reporting durations (Supplementary Figure E4) which may have led to an underestimate of the effect of smoking on the development of symptoms.

Among Standard Users, current smokers were more likely to develop the classic triad of symptoms suggesting a diagnosis of COVID-19 *(i.e*., fever, persistent cough and shortness of breath; OR[95% CI]=1.14[1.10 to1.18]; p=5.81×10^-13^) or to meet a higher threshold of symptom burden, than non-smokers (greater than 5 symptoms OR=1.29[1.26 to 1.31]; p=8.00×10^-187^; greater than 10 symptoms OR=1.50[1.42 to 1.58]; p=3.46×10^-48^) (**Figure 1**).

**Figure 1.**
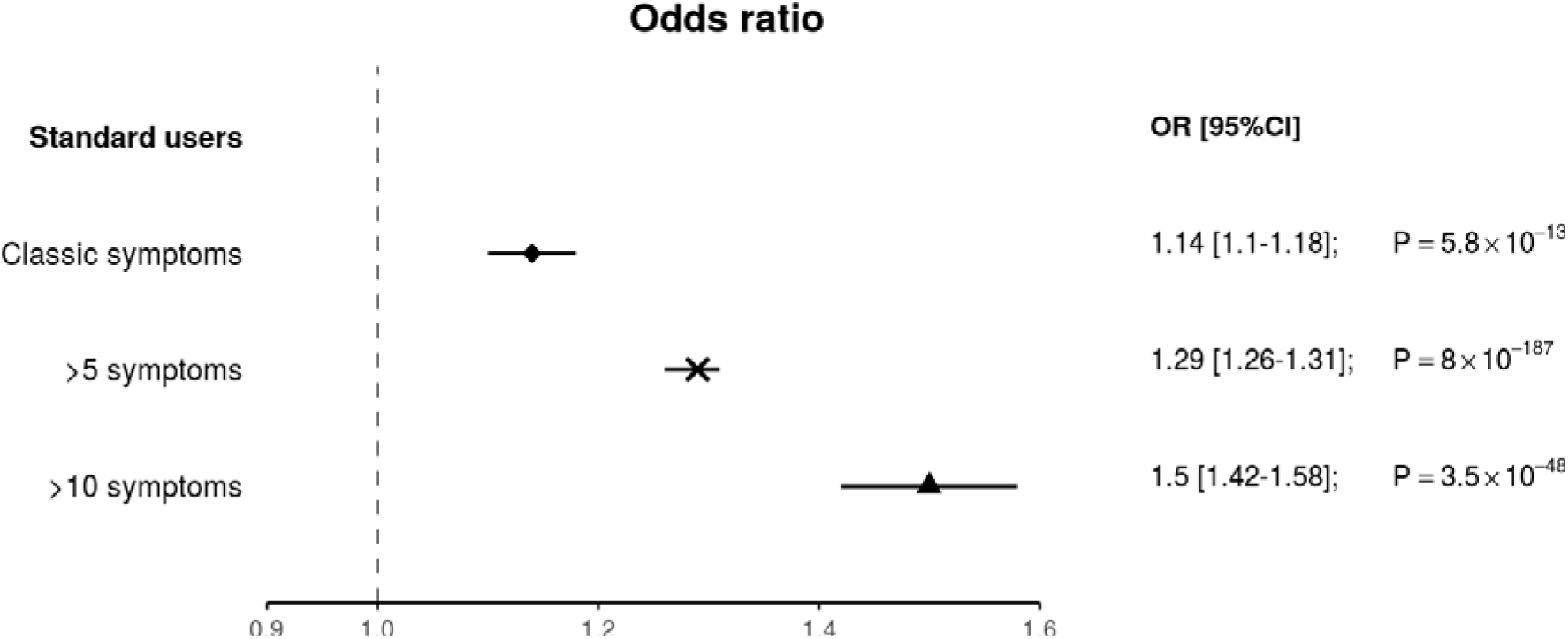
Effect of current smoking on risk of presenting with COVID-19 symptoms. The plot shows the OR [95% CI] for smokers from the Standard Users category of presenting with COVID-19 classic symptoms (i.e., all three of cough, fever and breathlessness) or a higher symptom burden (i.e., more than five or more than ten symptoms).

Smoking was strongly associated with having most COVID-19 symptoms among Standard Users and SC2S (p< 1.14×10^-3^; see **Methods; Table 2**). Of note current smokers testing positive for SARS-CoV-2 were more likely to develop delirium (OR[95% CI]=1.83 [1.47 to 2.27]; p=4.38×10^-8^) and abdominal pain (OR=1.43 [1.15 to 1.77]; p=1.07×10^-3^). Smoking was also associated with higher symptom burden in SC2S and, more importantly, SC2P; (OR [95%CI] of presenting with more than 10 symptoms 1.33 [1.22-1.45]; p=1.91×10^-10^ and 1.42 [1.09 to1.83]; p=7.13×10^-3^, respectively).

Among subjects who reported not feeling well, the incidence of all COVID-19 related symptoms, with the exception of delirium and sore throat, was higher in the SC2P group as compared to the other categories (Chi-squared test p<1.27×10^-6^; **Figure 2**). We did not observe any specific pattern of co-occurring symptoms distinguishing smokers from non-smokers, with symptom correlation matrices showing significant inter-correlation in all four user categories (lowest Mantel’s test *r^2^* = 0.54; p = 0.001; **Figure 3**).

**Table 2.** OR for individual symptoms in current smokers vs non-smokers. For each test group, the table reports the OR [95% CI] and p-value of the association between smoking status and individual COVID-19 symptoms, adjusted for age, sex and BMI. Associations passing Bonferroni correction for multiple testing (p<1.14×10^-3^) are highlighted in bold.

|  | Standard Users<br>n= 2,221,088 |  | COVID-19<br>self-reported<br>n=157,406 |  | SARS-CoV-2<br>Tested –<br>n=16,759 |  | SARS-CoV-2<br>Tested +<br>n=7,123 |  |
| --- | --- | --- | --- | --- | --- | --- | --- | --- |
| Symptom | OR [95% CI] | P | OR [95% CI] | P | OR [95% CI] | P | OR [95% CI] | P |
| Abdominal pain | **1.35 [1.33-1.38]** | **$8.35 \times 10^{-198}$** | **1.43 [1.37-1.50]** | **$7.21 \times 10^{-57}$** | **1.33 [1.13-1.56]** | **$6.94 \times 10^{-4}$** | 1.43 [1.15-1.77] | **$1.07 \times 10^{-3}$** |
| Chest pain | **1.16 [1.14-1.18]** | **$1.42 \times 10^{-63}$** | **1.10 [1.06-1.14]** | **$4.04 \times 10^{-6}$** | 1.06 [0.92-1.23] | $4.06 \times 10^{-1}$ | 1.31 [1.07-1.62] | $1.08 \times 10^{-2}$ |
| Delirium | **1.22 [1.20-1.25]** | **$2.12 \times 10^{-93}$** | **1.26 [1.20-1.31]** | **$1.08 \times 10^{-23}$** | **1.60 [1.34-1.89]** | **$8.09 \times 10^{-8}$** | **1.83 [1.47-2.27]** | **$4.38 \times 10^{-8}$** |
| Diarrhoea | **1.31 [1.29-1.33]** | **$3.94 \times 10^{-210}$** | **1.28 [1.23-1.34]** | **$5.59 \times 10^{-29}$** | **1.31 [1.12-1.53]** | **$5.55 \times 10^{-4}$** | 1.14 [0.92-1.40] | $2.34 \times 10^{-1}$ |
| Fatigue | **1.06 [1.05-1.08]** | **$1.75 \times 10^{-15}$** | 0.94 [0.90-0.99] | $1.40 \times 10^{-2}$ | 1.00 [0.84-1.18] | $9.55 \times 10^{-1}$ | 0.78 [0.57-1.10] | $1.46 \times 10^{-1}$ |
| Fever | 1.01 [0.98-1.03] | $6.37 \times 10^{-1}$ | 1.01 [0.96-1.06] | $8.42 \times 10^{-1}$ | 0.87 [0.74-1.02] | $8.79 \times 10^{-2}$ | 0.86 [0.70-1.06] | $1.54 \times 10^{-1}$ |
| Headache | **1.10 [1.08-1.12]** | **$1.91 \times 10^{-25}$** | **1.30 [1.24-1.36]** | **$2.77 \times 10^{-25}$** | 1.03 [0.87-1.22] | $7.23 \times 10^{-1}$ | 1.08 [0.83-1.41] | $5.89 \times 10^{-1}$ |
| Hoarse voice | **1.06 [1.04-1.08]** | **$8.38 \times 10^{-8}$** | 1.06 [1.01-1.10] | $1.04 \times 10^{-2}$ | 0.95 [0.81-1.12] | $5.48 \times 10^{-1}$ | 1.19 [0.97-1.46] | $1.02 \times 10^{-1}$ |
| Anosmia | **1.07 [1.05-1.09]** | **$6.98 \times 10^{-11}$** | **0.88 [0.85-0.92]** | **$4.82 \times 10^{-9}$** | 1.18 [0.99-1.39] | $5.55 \times 10^{-2}$ | 0.93 [0.74-1.17] | $5.32 \times 10^{-1}$ |
| Persistent cough | **1.18 [1.16-1.20]** | **$8.28 \times 10^{-90}$** | 0.99 [0.95-1.03] | $5.31 \times 10^{-1}$ | 0.88 [0.76-1.01] | $7.27 \times 10^{-2}$ | 0.73 [0.59-0.90] | $2.96 \times 10^{-3}$ |
| Shortness of breath | **1.16 [1.14-1.18]** | **$3.34 \times 10^{-71}$** | **1.09 [1.05-1.14]** | **$1.40 \times 10^{-5}$** | 1.11 [0.96-1.29] | $1.40 \times 10^{-1}$ | 1.35 [1.09-1.68] | $6.53 \times 10^{-3}$ |
| Skipped meals | **1.76 [1.73-1.79]** | **$P < 2.23 \times 10^{-308}$** | **1.52 [1.46-1.59]** | **$4.78 \times 10^{-88}$** | **1.46 [1.26-1.70]** | **$6.92 \times 10^{-7}$** | 1.31 [1.06-1.61] | $1.15 \times 10^{-2}$ |
| Sore throat | **0.93 [0.91-0.95]** | **$2.04 \times 10^{-14}$** | **1.17 [1.12-1.23]** | **$2.00 \times 10^{-11}$** | 0.90 [0.77-1.06] | $2.12 \times 10^{-1}$ | 1.32 [1.06-1.65] | $1.30 \times 10^{-2}$ |
| Unusual muscle pains | **1.47 [1.43-1.52]** | **$2.41 \times 10^{-151}$** | **1.46 [1.37-1.55]** | **$8.90 \times 10^{-30}$** | 1.24 [1.02-1.50] | $3.18 \times 10^{-2}$ | 1.19 [0.94-1.51] | $1.40 \times 10^{-1}$ |

**Figure 2.**
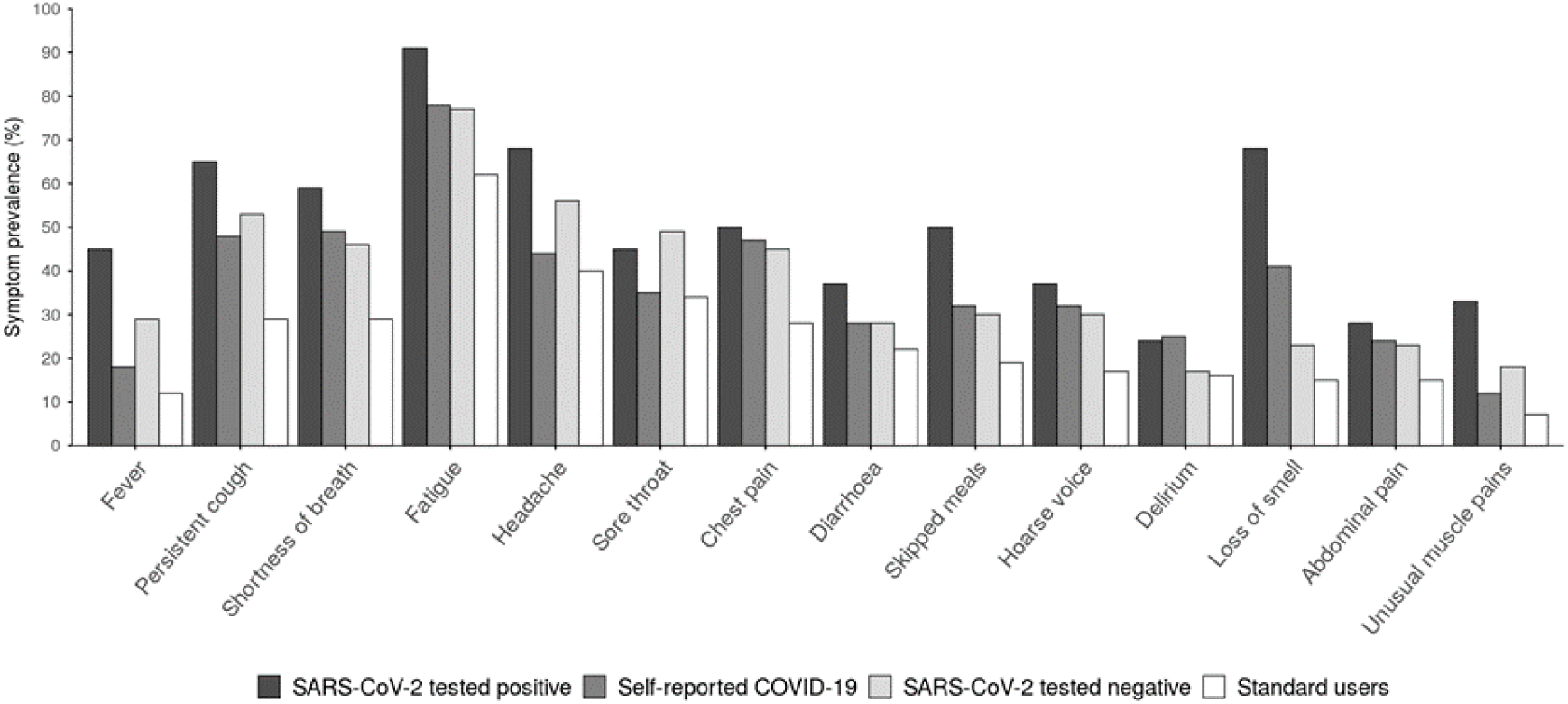
COVID-19 symptom distribution. Bar plot showing the proportion of individuals reporting COVID-19 symptoms within user categories. All symptoms except delirium and sore throat were more frequently observed among subjects who reported testing positive for SARS-CoV-2 (Chi-squared p<1.27×10^−6^).

**Figure 3.**
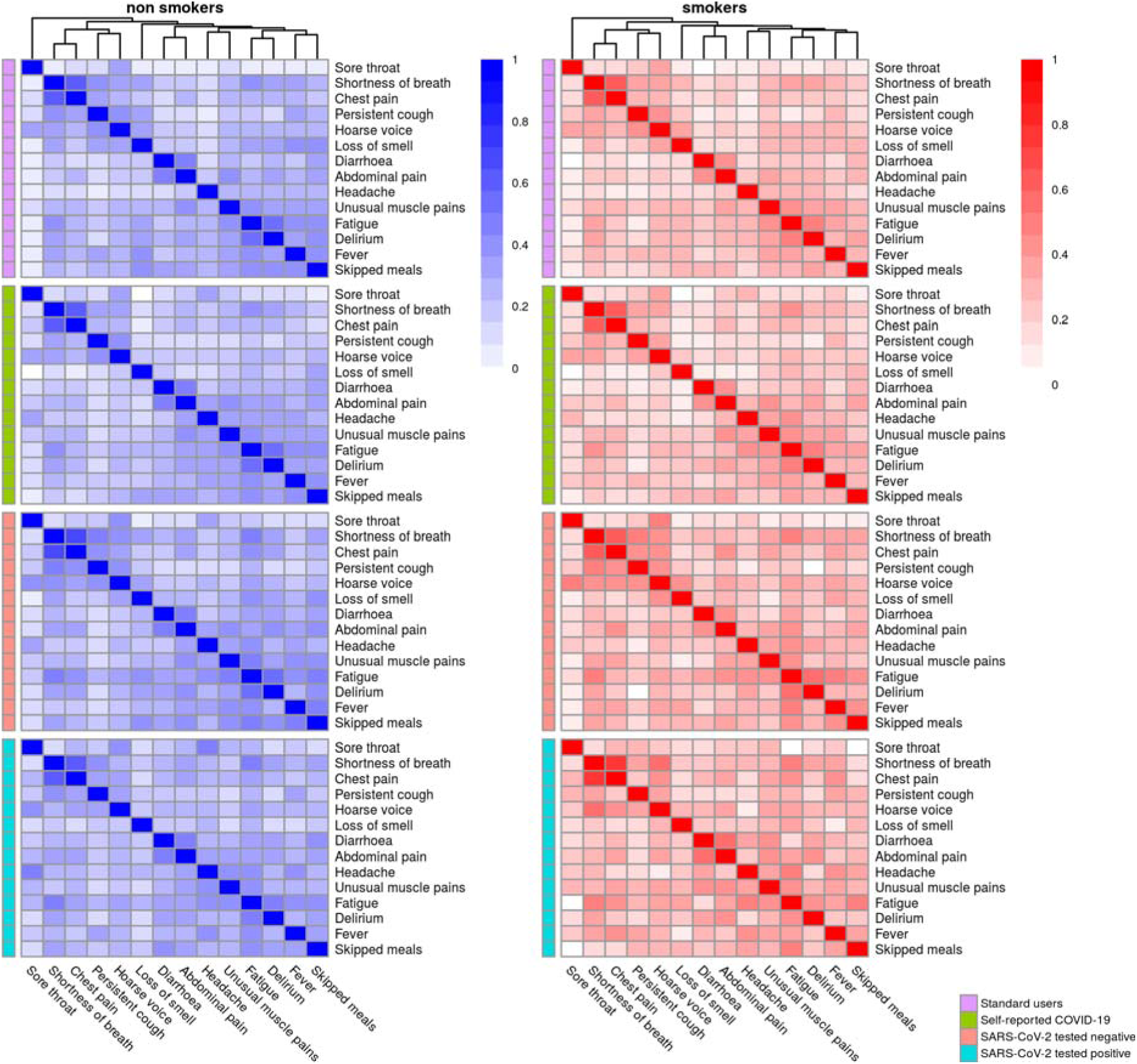
Correlation of symptoms in smokers and non-smokers. For each user category, the heatmaps represent the polychoric correlation matrices between COVID-19 symptoms according to smoking status (non-smokers in blue, and smokers in red).

To explore differences in COVID-19 disease course between smokers and non-smokers, we used the number of reported COVID-19 symptoms as a proxy for disease severity, with the assumption that a higher symptom burden was suggestive of more severe disease. Indeed, individuals reporting hospital attendance reported experiencing a higher number of co-occurring symptoms than those who did not attend hospital (OR 1.32[1.31 to 1.33]; P<2.22 ×10^-16^).

We observed a shift in the symptom burden distribution towards an increasing number of symptoms across groups, with the lowest average symptom burden in the Standard Users group, followed the by the self-assessed COVID-19 (SC2S) and by the tested negative (SC2N) categories, and the highest average symptom burden in the tested positive (SC2P) group (Wilcoxon’s test p< 2.22×10^-16^; Figure 3). Of particular interest, smoking was associated with increased overall COVID-19 symptom burden, in all categories (Standard Users *β*=2.93, SE=0.07, p<2.22×10^-16^; SC2S *β*=3.02, SE=0.21, p<2.22×10^-16^; SC2P *β*=3.28, SE=1.10, p=2.79×10^-3^; SC2N *β*=1.95, SE=0.78, p=1.22×10^-2^; **Figure 4**).

**Figure 4.**
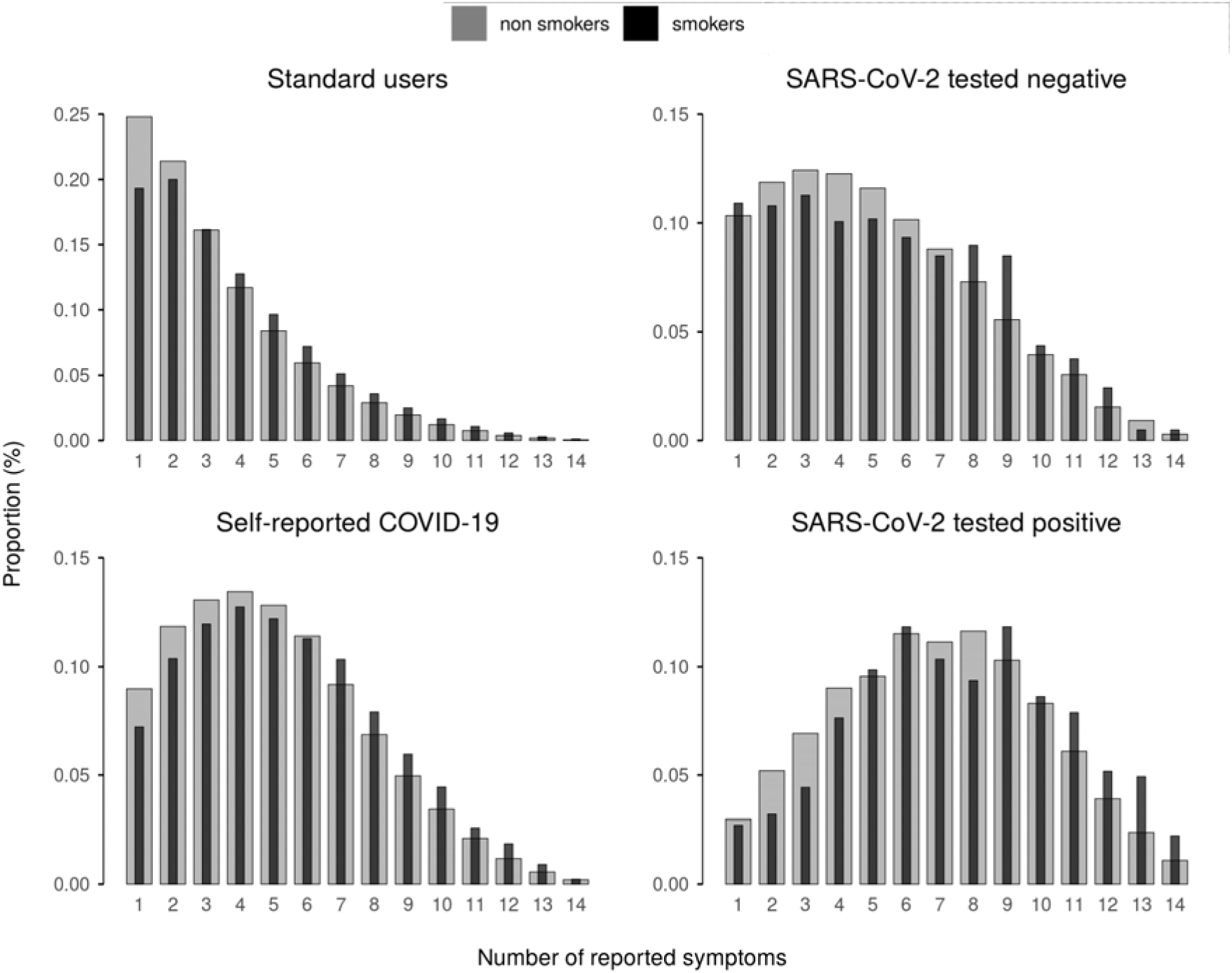
Effect of current smoking on COVID-19 symptom burden. For each user category, the bar plot shows the relative abundance of individuals by total number of COVID-19 symptoms reported, among non-smokers (grey) and smokers (black). Current smoking was associated with increased overall COVID-19 symptom burden in all categories.

### Characteristics of SARS-CoV-2 tested individuals

As expected, the group who reported that they had undergone testing (n=26,918 1.1% of app users) differed in a number of ways from the general study population (**Table 1**). In particular, 20% of those tested were healthcare workers (HCW) compared to 3.4% in the whole population and 31% of those testing positive were HCW. Smoking rates among tested individuals were lower compared to all users of the app (8.9% vs 11.0%). Smoking rates were lower in those who had tested positive for SARS-CoV-2 compared to those testing negative (7.4% vs 9.3%), as were rates of cancer, heart disease and lung disease. Although we observed an apparently protective effect, in the tested sub-population, of smoking on the risk of having a positive PCR for SARS-CoV-2 infection (OR [95% CI] 0.73 [0.65 to 0.81]; p=9.49×10^-9^), we also found that symptom burden was higher in smokers who tested positive for SARS-CoV-2 as compared to non-smokers. In addition, current smokers who tested positive had a higher risk of attending hospital due to COVID-19 (OR 2.11 [1.41 to 3.11] (p=1.99×10^-4^)). This association remained significant when subjects reporting the presence of comorbidities were removed from the analysis (OR 1.87 [1.15 to 2.95]; p=9.02×10^-3^).

### *ACE2* expression in GTEx lung and adipose tissue

We examined *ACE2* expression in publicly-available gene expression data measured by RNASeq in the GTEx project^24^. In this sample, median *ACE2* gene expression in subcutaneous adipose tissue was higher than in lung samples (**Supplementary figure E2**). *ACE2* expression levels were positively correlated between adipose and lung tissue from the same subjects (Spearman’s rho=0.25; p=1.57×10^-7^; **Supplementary Figure E3**).

### Association of *ACE2* expression with smoking in TwinsUK

We explored the relationship between *ACE2* expression in adipose tissue and smoking status in gene expression data measured by RNASeq in female twins from the TwinsUK cohort^1^. In this sample, smokers were found to have reduced *ACE2* expression in adipose tissue compared to non-smokers (Beta(SE)= −0.395(0.149); p=7.01×10^-3^). This association was robust to sensitivity analyses excluding subjects with type 2 diabetes, and to the inclusion of past smokers in the dataset (**Figure 5**) (**Supplementary Table E1**).

**Figure 5.**
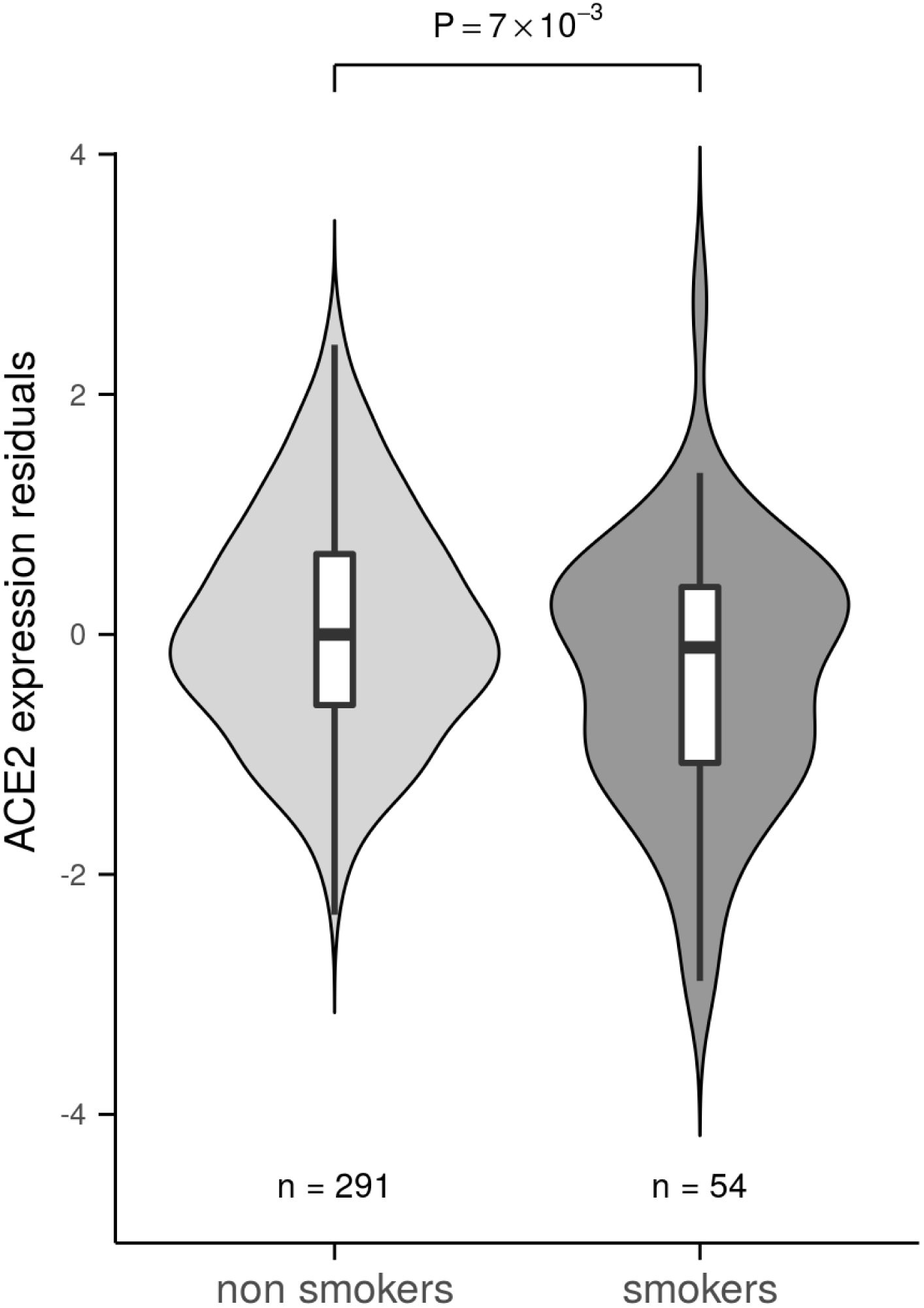
Association of smoking status with *ACE2* expression in adipose tissue. Among female participants in the TwinsUK study, smokers were found to have reduced *ACE2* expression in adipose tissue compared to non-smokers (p=7.01×10^−3^)).

## DISCUSSION

The main finding of this study from a large prospective population cohort was that current smoking was associated with a substantially increased risk of developing symptoms suggestive of COVID-19. In addition, current smoking was associated with a greater symptom burden, indicating an impact on disease severity and smokers testing positive for SARS-CoV-2 were more likely to attend hospital. *ACE2* expression in adipose tissue was noted to be lower which may have an impact on mechanisms of disease severity.

### Significance of findings

The COVID-19 pandemic spreads continues to threaten to overwhelm the capacity of health systems, especially as communities prepare for a release from social distancing after the first wave of the pandemic. There is a need for strategies to mitigate this, which includes preventing or delaying transmission of the virus to “flatten the curve”, as well as any measures that might reduce the severity of the condition. In addition, steps should be taken to increase health system capacity and reduce health system demand from other sources. The present data suggest that smoking cessation should be considered as an element in strategies to address COVID-19, as smoking increases both the likelihood of symptomatic disease defined according to the presence of “classic” symptoms of fever, cough and breathlessness and the severity of disease, defined based on the number of symptoms. Combined with this, a reduction in population smoking rates is likely to reduce the health system burden from other conditions that require hospitalisation such as acute vascular events and exacerbations of lung disease^8^ as well as improving resilience by reducing overall sickness absence among key-workers^26^.

In addition to an increase in individual susceptibility to developing COVID-19 following viral infection, smoking might also be expected to increase the risk of infection because of the repeated hand to mouth movements that smoking entails. The increase in severity that we found associated with smoking suggests that the whole curve of disease is shifted as opposed to just the chance of infection. Importantly, although the number and degree of symptoms was higher in smokers the pattern of symptoms did not differ, indicating that the phenomenon that we observed was not caused by the presence of symptoms related directly to smoking itself.

We found that adipose tissue *ACE2* expression levels were reduced in smokers compared to non-smokers in TwinsUK. The contribution of *ACE2* expression levels to Sars-CoV-2 infection and disease severity is a matter of intense interest^27^. It has been hypothesised that ACE2 internalisation after viral infection causes a reduction in ACE2 activity leading to increased Angiotensin II levels which contribute to the widespread endothelial damage, microthrombosis and coagulopathy observed in severe COVID-19^1,2^. It has also been hypothesised that reduced ACE2 levels may offer some protection against initial infection, but once infected, as is highly likely to occur at some stage in a pandemic setting, low ACE2 levels may contribute to increased disease severity^28^. Our data from TwinsUK showing that *ACE2* expression is lower in adipose tissue of smokers may explain some of the increased risk of systemic disease in smokers with COVID-19. The GTEx data indicate a weak correlation between adipose and lung *ACE2* expression. This could suggest that ACE2 expression may also be reduced in smokers’ lungs. However, there have also been recent studies showing the opposite effect^14-16^. To further complicate matters, single cell-type data suggest that in smokers *ACE2* is upregulated in epithelial cells but down-regulated in Type 2 alveolar cells^18^.

Data were also available for a limited number of individuals who reported the results of PCR tests performed to identify the presence of SARs-CoV-2. Only 1% of the study population had undergone testing and this group was heavily selected because of testing policies which focused on healthcare workers and others interacting with healthcare. As a consequence, extrapolation from this group to population risk is inappropriate. Smoking prevalence was lower in the tested group, 8.9% vs 11.0%, both values substantially below the UK population average and smoking prevalence was lower in those testing positive than those testing negative (7.4% vs 9.3%). This raises several possibilities. Firstly, it may be that smoking does reduce susceptibility to infection with SARS-CoV-2, though the likelihood of developing symptoms and severity of disease in current smokers remains higher, according both to the whole population data and the fact that smokers testing positive were more likely to report hospital attendance. Data from a multinational study of 8,910 hospitalised COVID-19 patients indicate that smokers experience more severe disease and have worse outcomes, including an increased risk of death (9.4%, vs. 5.6% among former smokers or non-smokers; OR [95% CI] 1.79 [1.29 to 2.47])^4^. Alternatively, our finding may represent sampling bias. Among those tested, a negative PCR result was also associated with a higher rate of heart disease, cancer and respiratory disease, all factors associated with worse outcomes in people with COVID-19 but unlikely to be protective against infection. The use of swab testing for PCR diagnosis of viral infection has limitations with a significant false negative rate. It is possible that non-random differences in the way that tests were conducted (e.g. nasopharyngeal or oropharyngeal swabs) or the temporal relation of testing to the development of symptoms (likely more rapid for healthcare workers for example) could have influenced results^29,30^. In due course, the question of relative infectivity may be answered based either on surveillance screening or widespread antibody testing.

### Methodological issues

There are some limitations in what can be inferred from these data. The study uses the development of symptoms to define COVID-19, but it is possible that some of these represented conditions other than COVID-19. Behavioural factors or individual prior expectations may have played a role. Thus smokers may have felt themselves to be at a higher risk of developing COVID-19 making them more likely to report symptoms, thus exaggerating the apparent impact of smoking. Alternatively, non-smokers being in general healthier, might be expected to be more sensitive to changes in their wellbeing making them more likely to report symptoms, thus underestimating the impact of smoking. Participants in the study were self-selected, having chosen to register to use the app. In addition the app was only available on a smartphone raising issues of digital access and digital literacy, so caution is needed about extrapolation to the entire population. Smoking rates were also lower in the study than in the general population and there were relatively fewer people in the older age groups who are most susceptible to the condition.

Our analyses of *ACE2* expression in adipose tissue in TwinsUK were limited by the small sample size of subjects with smoking data available. The expression dataset is also a female-only sample, which, given the sex differences observed in COVID-19 disease progression, limits inferences that can be drawn. Furthermore, previously noted differences in *ACE2* expression across tissues, and resultant associations, make extrapolation from adipose tissue challenging.

### Conclusion

At current trajectories, smoking will kill around one billion people in the 21^st^ century.^31^ Evidence that current smoking increases individual and thus health system risk from COVID-19 is a strong argument for governments to accelerate rather than pause measures to deliver on tobacco control plans.^32^ These should include options such as polluter-pays levies on tobacco transnationals that will continue, through the pandemic, to be highly profitable as well as enormously damaging^33^.

Our results provide compelling evidence for an association between current smoking and individual risk from COVID-19, including symptom burden and risk of attending hospital. Smoking cessation should be incorporated into public health campaigns and other efforts to address the COVID-19 pandemic.

## Data Availability

Anonymised research data will be shared with third parties via the centre for Health Data Research UK (HDRUK.ac.uk). Data updates can be found on https://COVID.joinzoe.com TwinsUK RNASeq gene expression data used in this study are available under EGA accession number EGAS00001000805.

## DATA SHARING

Anonymised research data will be shared with third parties via the centre for Health Data Research UK (HDRUK.ac.uk). Data updates can be found on https://COVID.joinzoe.com

TwinsUK RNASeq gene expression data used in this study are available under EGA accession number EGAS00001000805.

## CONTRIBUTORS

NSH, CS and MF conceived the analysis which was developed with input from all authors. Symptom data cleaning and analyses were performed by BM, MM, SO, NR and MF and gene expression analysis by JSE-M. NSH, NR and MF produced the first draft of the paper to which all authors contributed. All authors have reviewed and approved the final version. MF is the guarantor.

## DECLARATION OF INTERESTS

NSH is Chair of Action on Smoking and Health and Medical Director of The British Lung Foundation. TDS is a consultant to Zoe Global Ltd (Zoe) who developed the app. J W and RD are or employees of Zoe Global Limited. Other authors have no conflict of interest to declare.

## ACKNOWLEDGEMENTS

This work was supported by Zoe Global Limited as well as grants from the Wellcome Trust (212904/Z/18/Z) and the Medical Research Council (MRC)/British Heart Foundation Ancestry and Biological Informative Markers for Stratification of Hypertension (AIMHY; MR/M016560/1).

TwinsUK is funded by the Wellcome Trust, Medical Research Council, European Union, Chronic Disease Research Foundation (CDRF), Zoe Global Ltd and the National Institute for Health Research (NIHR)-funded BioResource, Clinical Research Facility and Biomedical Research Centre based at Guy’s and St Thomas’ NHS Foundation Trust in partnership with King’s College London. JSES, TDS and KSS acknowledge support from the Medical Research Council (MR/M004422/1).

The authors would like to thank members of the public for taking the time to enter their information into the ZOE COVID symptom study app in order to improve understanding of and response to the COVID-19 pandemic.

## TRANSPARENCY DECLARATION

Mario Falchi, the manuscript’s guarantor affirms that the manuscript is an honest, accurate, and transparent account of the study being reported; that no important aspects of the study have been omitted; and that any discrepancies from the study as planned (and, if relevant, registered) have been explained.

## Notes

### Clinical Trial

n/a

